# Brainstem and cerebellar volume loss and the associated clinical features in Progressive Supranuclear Palsy

**DOI:** 10.1101/2025.07.06.25330975

**Authors:** Chloe Spiegel, Timothy P. Siejka, Cassandra Marotta, Josh JY. Lee, Kelly Bertram, Terence J. O’Brien, Meng Law, Lucy Vivash, Ian H. Harding

## Abstract

**INTRODUCTION:** Progressive Supranuclear Palsy (PSP) is a neurodegenerative tauopathy. Tau pathology predominates in the basal ganglia and midbrain, with spread to other brain regions. Involvement of the cerebellum remains equivocal, but is increasingly recognized in the pathophysiology of neurological diseases. We hypothesize that volume loss is appreciable in cerebellum alongside brainstem regions.

**METHODS:** In this cross-sectional observational study volumetric brainstem and cerebellar subsegmentation of T1-weighted magnetic resonance imaging (MRI) was performed in 37 adults with PSP. Group-level comparisons were made with 38 adults with Parkinson’s disease (PD) and 35 healthy control (HC) subjects. Regional volumes in the PSP cohort were correlated against disease severity and cognition.

**RESULTS:** Compared with HC, the midbrain, corpus medullare and flocculonodular lobe were smaller in PSP (d=0.90-1.2). Compared with PD, the midbrain, pons and superior cerebellar peduncle (SCP) were smaller in PSP (d=0.82-1.9). There was a positive correlation between the frontal assessment battery (FAB) and volume of the superior (r=0.50) and inferior (r=0.48) cerebellar posterior lobes. The PSP rating scale also correlated with SCP (r=-0.58) and midbrain (r=-0.52) volume.

**CONCLUSION:** Additional regions of brainstem and cerebellar volume loss, alongside midbrain atrophy, were observed in PSP. The reported clinico-radiologic correlations suggest a role of the cerebellum in cognitive dysfunction. These findings indicate that the cerebellum is not spared in PSP, and support further work to understand the temporal course of cerebellar and cerebellar connectivity changes relative to other brain regions.

## Introduction

Progressive Supranuclear Palsy (PSP) is a rare atypical parkinsonian disorder typically characterised by supranuclear gaze palsy, axial rigidity and postural instability. It is a “4-repeat tauopathy” with predominance of tau-related pathology in the basal ganglia and midbrain^1^. From this epicentre, tau is postulated to spread rostrally and caudally over time, affecting numerous other regions heterogeneously across PSP variants^1^. In the most well recognised variant, PSP-Richardson’s syndrome (PSP-RS), typical oculomotor, axial and postural features are usually seen alongside non-motor features, in particular cognitive dysfunction. With disease progression, variable degrees of dysfunction are observed in motor and non-motor domains. Moreover, variant presentations are increasingly recognised, often attributed to early predominant regions of neurodegeneration (beyond the midbrain). For example, frontal or parietal neurodegeneration may result in ‘cortical’ phenotypes, such a PSP-Frontal (PSP-F) or PSP-Corticobasal syndrome (PSP-CBS), whereas a striatal predominance may be associated with PSP-parkinsonism (PSP-P)^1,2^.

Despite increasing recognition of variable tau spread and clinical heterogeneity in PSP, neuroimaging studies have primarily focused on measures of midbrain atrophy^3^. Whilst this remains an important clinical and research focus, and can support a PSP diagnosis, understanding the profile of neurodegeneration across the whole brain is critical to disentangling symptom evolution and variability. A growing body of work using MRI and PET image analysis in PSP cohorts has described changes in other supratentorial regions, particularly frontal cortex^3^. The involvement of the cerebellum in PSP remains more equivocal, despite this structure frequently being affected by tau pathology at autopsy^1,4^. While some MRI studies have reported significant cerebellar atrophy^5–8^, noting associations with cognition^9,10^, gait dysfunction^5,10^, and verbal fluency^10,11^, these are inconsistent and the specific morphology underlying these potential associations is not well understood. Moreover, there are conflicting reports from meta-analyses regarding the spatial profile of cerebellar grey matter alterations in PSP^12–15^. Nonetheless, a closer look at the infratentorial structures is warranted, particularly as cerebellar dysfunction is increasingly recognised to contribute to a broad range of clinical manifestations in diseases previously thought to spare the cerebellum^16–18^.

Taken together, current evidence supports a hypothesis that variable involvement of the cerebellum and other subtentorial structures across individuals with PSP may help explain the significant variability in symptom manifestation. The aim of this study is to characterise the patterns of atrophy in the brainstem and cerebellum in PSP using MRI segmentation and morphometry approaches that are optimised for subtentorial structures, and assess clinico-radiologic correlations with both motor and non-motor disease features.

## Methods

### 1. PROJECT DESIGN

Patients meeting the Movement Disorders Society (MDS)-PSP diagnostic criteria were recruited prospectively from the Movement Disorders clinics at Alfred Health, Melbourne, Australia. Allocations of PSP variants were made in accordance with the MDS-PSP diagnostic criteria by a movement disorders consultant neurologist. Sixteen PSP patients underwent clinical assessment and MRI via the Monash-Alfred atypical parkinsonian database ^19^. Twenty-two PSP patients underwent clinical assessment and MRI via a separate study conducted at multiple Australian sites. Clinical assessments common to both pathways included the PSP rating scale (PSPRS), Hoehn and Yahr (H&Y) scale, Frontal Assessment

Battery (FAB), and categorical fluency. The PSPRS is a dedicated PSP severity scale ranging from 0 (lowest severity) to 100 (greatest severity). There are six domains including historical features, mental, bulbar exam, ocular, limb and gait/midline examination. The H&Y scale ranges from 1 (unilateral parkinsonism with minimal disability) to 5 (confinement to bed or wheelchair unless aided), and the FAB comprises cognitive and behavioural tests, with scores ranging from 0 (impaired performance) to 18 (better performance). Categorical fluency is assessed over a minute, with more words per minute suggesting better performance^19^.

Demographic data including age, sex and disease duration, along with MRI scans, for thirty-nine PD and forty healthy control (HC) comparators were retrospectively collected from the Alfred Hospital, Melbourne, Australia (ethics project number 157/19). PD diagnoses were based on the medical records. The H&Y scale was estimated for PD patients based on the clinical records available at the time of MRI.

### 2. STANDARD PROTOCOL APPROVALS, REGISTRATIONS, AND PATIENT CONSENTS

Ethics approval was obtained from the Human Research Ethics Committee at the Alfred Hospital (*Monash-Alfred atypical parkinsonian database* project number 157/19; *SEL003* project number 594/20; ACTRN12600001254987). Written informed consent was obtained from all participants recruited.

### 3. MRI ACQUISITION AND PROCESSING

All PSP patients underwent a standardised brain MRI protocol on a 3-Tesla Siemens scanner, including a 3D T1-weighted Magnetization Prepared Rapid Gradient Echo (MPRAGE) sequence with isotropic 0.8mm (clinical trial) or 1.0mm voxels (atypical parkinsonian database). Retrospectively curated MRI data for the comparator groups largely ranged in resolution from isotropic 0.9mm to 1.25mm voxels.

The MPRAGE images were processed to generate regional brain volume estimates using Freesurfer 7.3.2, Fastsurfer 2.2.0, and the Computational Anatomy Toolbox (CAT12) version 1113 in SPM12 version 7487 for MATLAB 9.4 (R2018a). Volumes of the midbrain, pons, and medulla were calculated using the *brainstem segmentation* pipeline in Freesurfer; the estimated Total Intracranial Volume (eTIV) for each scan was also recorded. The volumes of thirty cerebellar regions were calculated using the *CerebNet* cerebellar segmentation pipeline in Fastsurfer^20^, and then consolidated into six regions of interest (ROI): the bilateral cerebellar white matter, anterior lobe (lobules I-V), superior posterior lobe (lobules VI-VII), inferior posterior lobe (lobule VIII-IX), flocculonodular lobe (lobule X), and the vermis. Brainstem and cerebellar ROI are shown in **Supplementary Figure 1**.

Voxel-based morphometry (VBM) was used to calculate volumes of the superior cerebellar peduncle (SCP), middle cerebellar peduncle (MCP), Inferior Cerebellar Peduncle (ICP) and the dentate region of the cerebellum. The T1-weighted scans were processed in CAT12 to segment the brain into grey matter, white matter, and CSF, and then nonlinearly register the white matter segment to standard MNI space. The Jacobian determinant of the registration was calculated and used to assign an estimated volume value to each voxel in MNI space, as per standard VBM approaches. The volume of each ROI was calculated by summing the values of all voxels within bilateral masks of the three peduncles provided by the John Hopkins University (JHU) atlas^21^, and for the dentate nuclei using the SUIT atlas^22^.

Individual differences in head size were accounted for by scaling all volume measures by eTIV prior to statistical analyses (i.e., all volumes are expressed as %eTIV). Freesurfer eTIV estimate was used for ROIs derived from the Freesurfer and Fastsurfer pipelines, while the CAT12 eTIV estimate was used for the VBM-based ROIs.

### 4. QUALITY CONTROL

MRIQC version 24.1.0 and CAT12 were used to assess scan quality based on automated metrics of image properties including signal-to-noise ratio (SNR) and Full Width at Half Maximum (FWHM). Two scans were subsequently removed due to excessive image smoothness (FWHM) associated with considerable motion artefact. Using the CAT12 image and preprocessing quality metrics, one additional scan was removed due to very poor resolution (D+ 67.37%), likely due to partial volume effects.

Segmentation quality was additionally assessed by comparing the eTIV estimates derived by Freesurfer and by CAT12. Although subtle differences are expected, these estimates should be highly concordant and correlated. We noted five cases of scans with significant discrepancy (>300cm^2^) in these measures, suggesting a processing error in at least one of the pipelines. As outcomes from both pipelines are used in our analyses, these scans were excluded. As an additional quality check, we compared the eTIVs across sex and disease groups. As expected, the eTIV was larger in males than females (Freesurfer mean eTIV in males 1608cm^2^ and females 1415cm^2^; CAT12 mean eTIV in males 1627cm^2^ and females 1433cm^2^, p<0.001). There were no significant differences across the disease groups for the CAT12 eTIV (PSP 1554cm^2^ HC 1503cm^2^ PD 1562^2^, p=0.197), however, the Freesurfer eTIV was noted to be significantly larger in the PSP group (PSP 1588cm^2^ HC 1475cm^2^ PD 1501cm^2^, p=0.013). All volume measures are scaled by eTIV at the individual level to account for this potential confound in all analyses.

### 5. STATISTICAL ANALYSIS

Statistical analysis was performed using Jamovi 2.3.28. Data were reported using descriptive statistics, expressed in percentages, measures of central tendency, and spread. Comparisons across multiple groups were made using one-way ANOVA and χ^2^ tests. Independent samples student’s t-tests were used for pairwise between-group comparisons. Clinico-radiologic correlations with PSPRS, FAB, and Category Fluency were determined using linear regression analysis with correction for age, sex, and time since diagnosis. Missing data were indicated, and excluded from relevant analyses. A Bonferroni corrected p-value of 0.0038 was considered significant for all analyses (raw p-value threshold of 0.05 corrected for 13 regions). Outcomes meeting uncorrected p<0.05 thresholds are also indicated.

## 6. DATA AVAILABILITY

Anonymized data from the Monash-Alfred Atypical Parkinsonian Database are available and may be requested by a qualified investigator. Data from the ongoing SEL003 study will not be available until its completion in 2026. Data sharing requires Human Research Ethics Committee approval and a signed agreement.

## Results

### 1. DEMOGRAPHICS AND CLINICAL FINDINGS

Demographic data across PSP, PD and HC groups are reported in **Table 1**. All three groups comprised similar age and sex distribution. Time since diagnosis was significantly shorter in the PSP group compared with PD (2.74 years in PSP vs 6.48 years in PD, p<0.001). The median Hoehn and Yahr score was significantly higher in the PSP group compared with PD (3.5 in PSP vs 2.0 in PD, p<0.001). Clinical findings in the PSP group are reported in **Table 2**, with a breakdown of the variants included. This cohort included 36 individuals with probable PSP, comprising 33 PSP-RS, 2 PSP-CBS, 1 PSP-P and 1 PSP-PGF variants. The individuals with PSP-P and PSP-PGF had a notably longer disease duration. Statistical comparisons between variants were not performed due to the small sample sizes.

**Table 1:** Demographics, disease duration and severity.

|  | PSP (n=37) | PD (n=38) | HC (n=35) | p value |
| --- | --- | --- | --- | --- |
| Age (Mean (SD)) | 68.32 (6.65) | 69.12 (11.17) | 65.23 (8.37) | 0.149<sup>a</sup> |
| Sex (Male %) | 21 (57%) | 22 (58%) | 18 (51%) | 0.841<sup>b</sup> |
| Time since diagnosis (Mean (SD)) | 2.74 (2.95) | 6.48 (5.87) | - | <0.001<sup>c</sup> |
| H&Y (Median (range)) | 3.5 (3-5) | 2.0 (1-4) | - | <0.001<sup>c</sup> |

Statistical analysis using <sup>a</sup> one-way ANOVA, <sup>b</sup> $\chi^2$ test, <sup>c</sup> student's t-test
PSP = Progressive Supranuclear Palsy, PD = Parkinson's disease, HC=Healthy Controls, H&Y = Hoehn and Yahr scale

**Table 2:** Clinical characteristics in PSP.

|  | *PSP (n=37)* | *PSP-RS (n=33)* | *PSP-P (n=1)* | *PSP-PGF (n=1)* | *PSP-CBS (n=2)* |
| --- | --- | --- | --- | --- | --- |
| Time since diagnosis (years) | 2.74 (2.95) | 2.39 (2.47) | 8.30 | 11.60 | 1.35 (0.35) |
| PSPRS | 36.8 (14.60) | 36.6 (12.78)<sup>a</sup> | 20 | 25 | 68 (0.00) |
| FAB | 13.3 (3.55) | 13.65 (3.21)<sup>a</sup> | 18 | 15 | 6.50 (2.12) |
| Categorical fluency | 11.6 (4.78) | 12.00 (4.14)<sup>a</sup> | 19 | 15 | 2.00 (2.82) |

Results reported as Mean (SD) for groups, <sup>a</sup> Data missing for one subject
PSP = Progressive Supranuclear Palsy, PSP-RS = Progressive Supranuclear Palsy-Richardson Syndrome, PSP-P = Progressive Supranuclear Palsy-Parkinsonism variant, PSP-PGF = Progressive Supranuclear Palsy-Progressive Gait Freezing variant, PSP-CBS = Progressive Supranuclear Palsy-Corticobasal Syndrome variant, PSPRS = Progressive Supranuclear Palsy Rating Scale, FAB = Frontal Assessment Battery

### 2. VOLUMETRIC BRAINSTEM AND CEREBELLAR FINDINGS

The results of comparisons using mean percentage volume of eTIV are reported in **Table 3** and data are shown in **Figure 1**.

**Table 3:** Comparisons of regional volumetric findings.

|  |  |  | PSP vs PD |  |  | PSP vs HC |  |  |
| --- | --- | --- | --- | --- | --- | --- | --- | --- |
|  |  | PSP (%) | PD (%) | P value | Effect Size | HC (%) | P value | Effect size |
| **Brain stem** | Midbrain | 0.340 (0.04) | 0.413 (0.37) | **<0.001\*\*** | 1.864 | 0.385 (0.03) | **<0.001\*\*** | 1.182 |
|  | Pons | 0.893 (0.13) | 1.005 (0.12) | **<0.001\*\*** | 0.924 | 0.950 (0.10) | 0.036* | 0.503 |
|  | Medulla | 0.280 (0.035) | 0.293 (0.03) | 0.098 | 0.387 | 0.282 (0.04) | 0.873 | 0.038 |
| **Cerebellar Peduncles** | SCP | 0.068 (0.01) | 0.075 (0.01) | **<0.001\*\*** | 0.817 | 0.071 (0.01) | 0.064 | 0.444 |
|  | MCP | 0.668 (0.09) | 0.693 (0.07) | 0.188 | 0.307 | 0.696 (0.06) | 0.129 | 0.362 |
|  | ICP | 0.083 (0.01) | 0.086 (0.01) | 0.145 | 0.341 | 0.086 (0.01) | 0.117 | 0.374 |
| **Deep CBLM** | Dentate region | 1.026 (0.18) | 1.073 (0.02) | 0.224 | 0.283 | 1.105 (0.10) | 0.024* | 0.543 |
|  | Corpus Medullare | 1.537 (0.21) | 1.645 (0.22) | 0.034* | 0.500 | 1.708 (0.16) | **<0.001\*\*** | 0.896 |
| **Cerebellar Grey Matter** | Anterior Lobe | 0.738 (0.10) | 0.798 (0.09) | 0.007 | 0.642 | 0.756 (0.10) | 0.425 | 0.189 |
|  | Superior Posterior Lobe | 3.404 (0.51) | 3.620 (0.46) | 0.059 | 0.443 | 3.628 (0.36) | 0.037* | 0.502 |
|  | Inferior Posterior Lobe | 1.440 (0.21) | 1.501 (0.17) | 0.169 | 0.321 | 1.526 (0.20) | 0.079 | 0.420 |
|  | Flocculonodular Lobe | 0.076 (0.01) | 0.084 (0.02) | 0.013* | 0.589 | 0.088 (0.01) | **<0.001\*\*** | 0.986 |
|  | Vermis | 0.322 (0.04) | 0.336 (0.04) | 0.150 | 0.336 | 0.348 (0.03) | 0.006* | 0.665 |

Regional volume as percentage of eTIV reported as Mean (SD) for each group
P values and Effect Sizes calculated using student's t- test and Cohen's d
\*p<0.05 \*\* p<0.0038 (Bonferroni correction)

**Figure 1.**
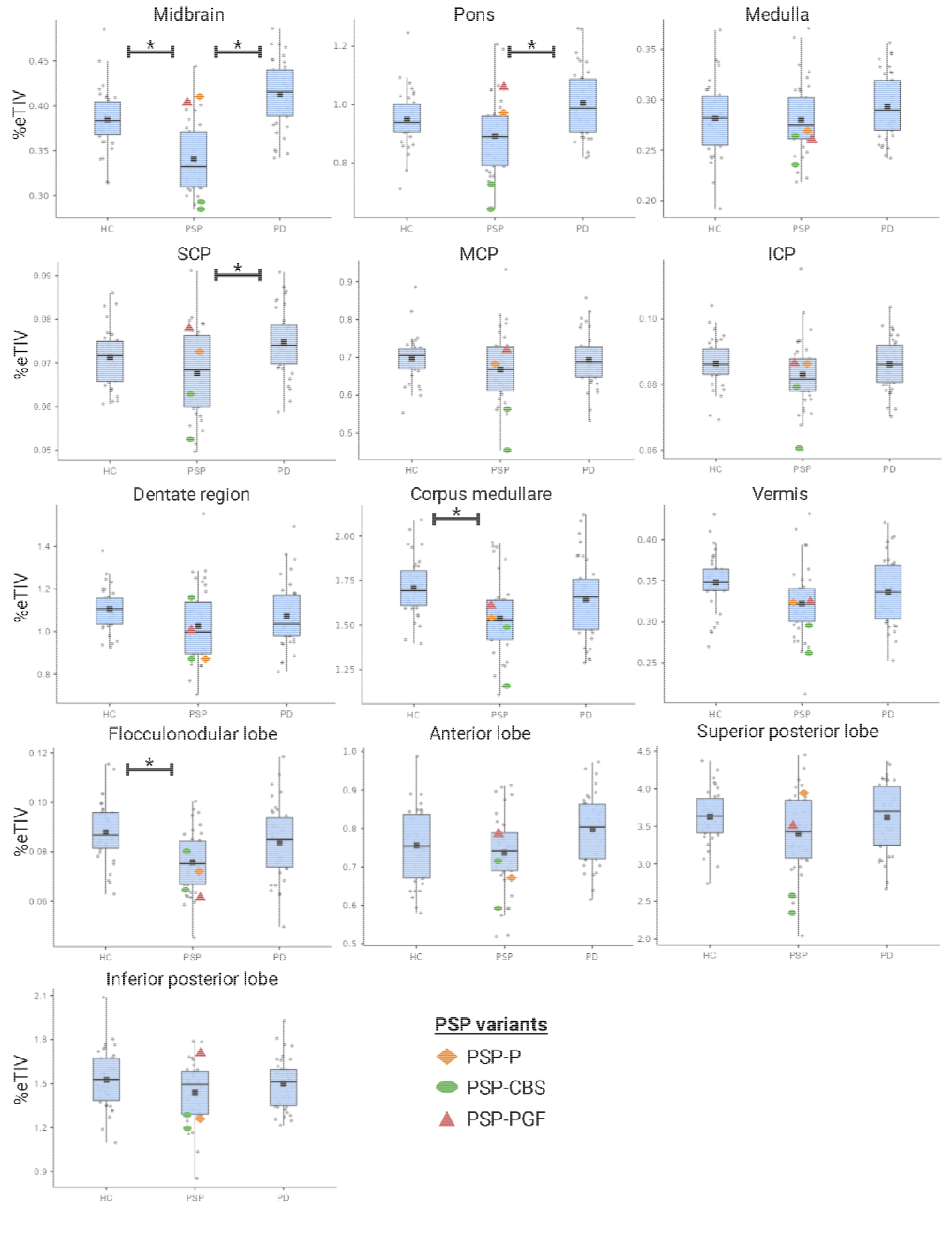
Box plots of regional volumetric comparisons with PSP variants. * p<0.0038 (Bonferroni correction) PSP = Progressive Supranuclear Palsy, PD = Parkinson’s Disease, HC = Healthy Controls, SCP = Superior Cerebellar Peduncles, MCP = Middle Cerebellar Peduncles, ICP = Inferior Cerebellar Peduncles, eTIV = estimated Total Intracranial Volume

#### PSP vs HC

Midbrain volume was significantly lower in PSP compared with HC with a very large effect size (d=1.2, p<0.001). A medium effect size group difference in pons volume was also observed (d=0.50, p=0.036), but did not survive correction for multiple comparisons. In the cerebellum, the corpus medullare (d=0.90, p<0.001) and flocculonodular lobe (d=0.99, p<0.001) were significantly reduced compared with HC with large effect sizes. Trends towards reduced volume with medium effect sizes were also observed in the dentate region (d=0.54, p=0.024), vermis (d=0.67, p=0.006), and superior posterior lobe (d=0.50, p=0.037), however these did not survive correction for multiple comparisons.

#### PSP vs PD

Compared with PD, midbrain (d=1.9, p<0.001), pons (d=0.92, p<0.001), and SCP (d=0.82, p<0.001) volumes were significantly lower in PSP. Medium effect size differences in the anterior lobe (d=0.64, p=0.007), cerebellar white matter (d=0.50, p=0.034), and flocculonodular lobe (d=0.59, p=0.013) were also observed, but did not survive correction. Analysis of the PSP-RS group alone, omitting the other rare variants, versus HC and PD revealed the same pattern of results with only minor deviations in effect size and significance estimates.

### 3. CLINICO-RADIOLOGIC CORRELATIONS

Findings for each clinical variable are reported in **Table 4** for the entire PSP group and for the PSP-RS only group.

**Table 4:**
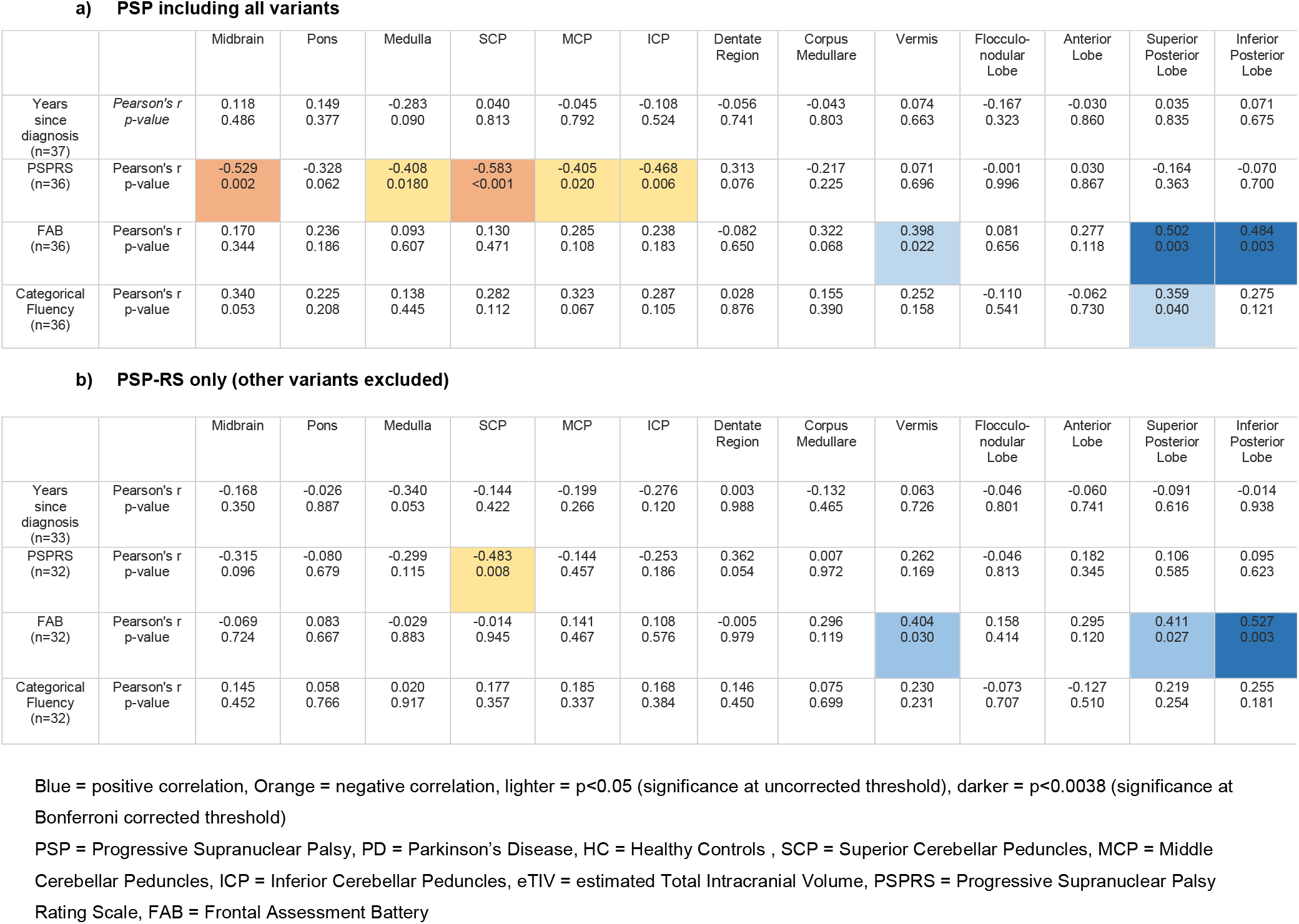
Clinico-radiologic correlations.

**a) PSP including all variants**
|  |  | Midbrain | Pons | Medulla | SCP | MCP | ICP | Dentate Region | Corpus Medullare | Vermis | Flocculo-nodular Lobe | Anterior Lobe | Superior Posterior Lobe | Inferior Posterior Lobe |
| --- | --- | --- | --- | --- | --- | --- | --- | --- | --- | --- | --- | --- | --- | --- |
| Years since diagnosis (n=37) | Pearson's r<br>p-value | 0.118<br>0.486 | 0.149<br>0.377 | -0.283<br>0.090 | 0.040<br>0.813 | -0.045<br>0.792 | -0.108<br>0.524 | -0.056<br>0.741 | -0.043<br>0.803 | 0.074<br>0.663 | -0.167<br>0.323 | -0.030<br>0.860 | 0.035<br>0.835 | 0.071<br>0.675 |
| PSPRS (n=36) | Pearson's r<br>p-value | -0.529<br>0.002 | -0.328<br>0.062 | -0.408<br>0.0180 | -0.583<br><0.001 | -0.405<br>0.020 | -0.468<br>0.006 | 0.313<br>0.076 | -0.217<br>0.225 | 0.071<br>0.696 | -0.001<br>0.996 | 0.030<br>0.867 | -0.164<br>0.363 | -0.070<br>0.700 |
| FAB (n=36) | Pearson's r<br>p-value | 0.170<br>0.344 | 0.236<br>0.186 | 0.093<br>0.607 | 0.130<br>0.471 | 0.285<br>0.108 | 0.238<br>0.183 | -0.082<br>0.650 | 0.322<br>0.068 | 0.398<br>0.022 | 0.081<br>0.656 | 0.277<br>0.118 | 0.502<br>0.003 | 0.484<br>0.003 |
| Categorical Fluency (n=36) | Pearson's r<br>p-value | 0.340<br>0.053 | 0.225<br>0.208 | 0.138<br>0.445 | 0.282<br>0.112 | 0.323<br>0.067 | 0.287<br>0.105 | 0.028<br>0.876 | 0.155<br>0.390 | 0.252<br>0.158 | -0.110<br>0.541 | -0.062<br>0.730 | 0.359<br>0.040 | 0.275<br>0.121 |

**b) PSP-RS only (other variants excluded)**
|  |  | Midbrain | Pons | Medulla | SCP | MCP | ICP | Dentate Region | Corpus Medullare | Vermis | Flocculo-nodular Lobe | Anterior Lobe | Superior Posterior Lobe | Inferior Posterior Lobe |
| --- | --- | --- | --- | --- | --- | --- | --- | --- | --- | --- | --- | --- | --- | --- |
| Years since diagnosis (n=33) | Pearson's r<br>p-value | -0.168<br>0.350 | -0.026<br>0.887 | -0.340<br>0.053 | -0.144<br>0.422 | -0.199<br>0.266 | -0.276<br>0.120 | 0.003<br>0.988 | -0.132<br>0.465 | 0.063<br>0.726 | -0.046<br>0.801 | -0.060<br>0.741 | -0.091<br>0.616 | -0.014<br>0.938 |
| PSPRS (n=32) | Pearson's r<br>p-value | -0.315<br>0.096 | -0.080<br>0.679 | -0.299<br>0.115 | -0.483<br>0.008 | -0.144<br>0.457 | -0.253<br>0.186 | 0.362<br>0.054 | 0.007<br>0.972 | 0.262<br>0.169 | -0.046<br>0.813 | 0.182<br>0.345 | 0.106<br>0.585 | 0.095<br>0.623 |
| FAB (n=32) | Pearson's r<br>p-value | -0.069<br>0.724 | 0.083<br>0.667 | -0.029<br>0.883 | -0.014<br>0.945 | 0.141<br>0.467 | 0.108<br>0.576 | -0.005<br>0.979 | 0.296<br>0.119 | 0.404<br>0.030 | 0.158<br>0.414 | 0.295<br>0.120 | 0.411<br>0.027 | 0.527<br>0.003 |
| Categorical Fluency (n=32) | Pearson's r<br>p-value | 0.145<br>0.452 | 0.058<br>0.766 | 0.020<br>0.917 | 0.177<br>0.357 | 0.185<br>0.337 | 0.168<br>0.384 | 0.146<br>0.450 | 0.075<br>0.699 | 0.230<br>0.231 | -0.073<br>0.707 | -0.127<br>0.510 | 0.219<br>0.254 | 0.255<br>0.181 |

Blue = positive correlation, Orange = negative correlation, lighter = $p < 0.05$ (significance at uncorrected threshold), darker = $p < 0.0038$ (significance at Bonferroni corrected threshold)
PSP = Progressive Supranuclear Palsy, PD = Parkinson's Disease, HC = Healthy Controls, SCP = Superior Cerebellar Peduncles, MCP = Middle Cerebellar Peduncles, ICP = Inferior Cerebellar Peduncles, eTIV = estimated Total Intracranial Volume, PSPRS = Progressive Supranuclear Palsy Rating Scale, FAB = Frontal Assessment Battery

#### Disease Duration

There were no regional volumes that significantly correlated with time since diagnosis. A separate analysis with only PSP-RS (variants excluded) also did not show any significant correlations.

#### Disease Severity

The PSPRS was observed to negatively correlate at corrected-level significance with midbrain (p=0.002, R=-0.529) and SCP (p<0.001, R=-0.583) volumes. Negative correlations between medulla (p=0.018 R=-0.408), MCP (p=0.020 R=-0.405) and ICP (p=0.006, R=-0.468) were also present at uncorrected significance. No significant correlations were observed with cerebellar ROIs. The analysis of PSP-RS only showed a similar negative correlation with SCP volume (p=0.008, R=-0.483) which did not meet the corrected significance threshold.

#### Cognition

There was a significant positive correlation between the FAB and cerebellar superior posterior (p=0.003 R=0.502) and inferior posterior (p=0.003, R=0.484) lobe volumes. Analysis of only PSP-RS showed the same relationship between the FAB and posterior lobe regions (inferior: p=0.003, R=0.527; superior: p=0.027, R=0.411), but which no longer reached corrected significance in the superior posterior region. A similar trend was noted between the FAB and the vermis volume (p=0.022, R=0.398), and between categorical fluency and cerebellar superior posterior lobe (p=0.040 R=0.359) volume. While the relationship between FAB and vermis (p=0.030, R=0.404) volume was comparable on the analysis of PSP-RS, no similar relationships were observed with categorical fluency.

## Discussion

In this study we have identified several infratentorial regions of volume loss beyond the midbrain in PSP. Of note, significant volume loss was seen in the pons and several cerebellar regions, alongside midbrain and SCP atrophy. Significant clinico-radiologic correlations were noted with disease severity and cognition. Importantly, while disease severity negatively correlated with brainstem and peduncle volumes, cognitive impairment was noted to positively correlate with the superior and inferior cerebellar posterior lobe volumes.

As expected, significant midbrain volume loss was detected in PSP compared with both Parkinson’s disease and healthy control groups, and this was correlated with disease severity. This result is consistent with prior volumetric studies in PSP patients^9,23,24^, and concordant with the known pathophysiology of this disease. Interestingly, there was no significant correlation with disease duration, which may indicate variable tau spread and neurodegeneration. Volume loss was also noted in the pons in our study, particularly in comparison with Parkinson’s disease. Despite relative sparing of the caudal brainstem regions in PSP, there have been several studies reporting significant pontine atrophy^8,25–27^, including a prior meta-analysis^28^. Similar to many prior studies^29–32^, we did not find significant volumetric differences in the medulla. These findings are consistent with the central involvement of the midbrain in PSP pathology, but also suggest more subtle changes in the pons which may be clinically relevant.

In the cerebellar peduncles, we report significant volume loss in the SCP relative to PD, although not in comparison with HC, and we show that SCP volume is associated with disease severity. The SCP predominantly carries efferent fibers that emerge from cerebellar dentate nuclei and form the first leg of the ascending dentato-thalamo-cortical pathway. SCP volume loss has been reported across many studies in PSP^24,25,28,31^, and appears to coincide with midbrain atrophy^24,33^. It is unexpected that the SCPs were not significantly smaller in the comparison with HC, however there was an apparent trend. There were no significant volumetric differences observed in the MCP and ICP, which is compatible with relative sparing of these structures. Intriguingly, despite this, the ICP volume was found to negatively correlate with disease severity, and a relationship with a similar effect size with MCP volume was also found, although fell outside of corrected-level significance. The MCP predominantly carries afferent fibres to the cerebellum, forming the second leg of the cortico-ponto-cerebellar tract, while the ICP predominantly carries afferent fibres from the spinal cord, vestibular system and lower brainstem to the cerebellum. The potential implications of damage to these structures would be wide-ranging, affecting motor and cognitive domains by impairing vital cerebro-cerebellar loops. Moreover, although not observed in our study, there are prior reports of volume loss in the MCPs and ICPs in PSP^33,34^. It is feasible that small volumetric changes in the relatively spared MCPs and ICPs contribute multifactorially to disease severity, alongside the more robust changes observed in the SCPs.

In the cerebellum, significant volume loss was noted in the corpus medullare in PSP relative to healthy controls, and Parkinson’s disease to a lesser extent. Though reports of cerebellar changes in PSP have been inconsistent, such findings are in-line with some prior volumetric studies^6–8,35^. It is important to note that the corpus medullare comprises both the cerebellar white matter and deep cerebellar nuclei, including the dentate nucleus. The dentate nuclei are the primary output hubs of the cerebellum, and give rise to the SCPs. Several tau PET studies have shown increased uptake in the dentate nucleus^24,32,36^ and diffusion imaging studies have shown microstructural impairments along this pathway in PSP^8,37^. We therefore examined the dentate region in isolation from the corpus medullare using a voxel-based approach. While the volume of the dentate nucleus region was significantly lower in PSP compared with healthy controls, this did not survive correction and was not seen in the comparison with Parkinson’s disease. This finding suggests that the volumetric difference observed in the corpus medullare is not entirely attributable to changes in the dentate nucleus, indicating the likelihood of white matter volume loss. This is consistent with reports of tau pathology in the cerebellar white matter at autopsy across multiple PSP variants^1,4^.

A consistent pattern of smaller volumes in cerebellar cortex regions was observed in the PSP cohort relative to both PD and healthy controls, but with varying magnitude across regions (d=0.2-1.0) and only sparsely meet our stringent criteria for statistical significance. Whilst these observations therefore need to be interpreted with caution, and must be confirmed in a larger study cohort, this pattern suggests a relatively subtle involvement of cerebellar cortex in PSP pathology. The clinical relevance of these observations are indicated by our finding that superior and inferior posterior lobe volumes correlated with cognitive functioning, assessed by the FAB, in the PSP cohort. Involvement of the cerebellar posterior lobe in PSP is further supported by a voxel based meta-analysis from 2017 which reported significant volume loss in crus I, crus II, lobule VIIb in the superior posterior lobe, as well as lobule IX in the inferior posterior lobe in PSP^13^. Moreover, functional MRI studies have revealed connectivity of the posterior lobe with several networks underpinning cognitive function, including the executive control network, dorsal and ventral attention networks, and the default mode network^38^.

Disruption of these networks is postulated to contribute to cognitive dysfunction in PSP, along with several other neurodegenerative disorders^13^. These findings align with the current understanding of cerebellar functional topography and highlight critical involvement of the cerebellum in non-motor PSP symptoms.

Taken together with variable reports of involvement of other cerebellar regions, including the anterior lobe and vermis, in PSP^8,11,12,35^, the current study provides exciting indications that emphasise the need to better characterise cerebellar alterations in PSP. Further studies of the cerebellum in association with other non-motor symptoms in PSP, such as affect, mood and language would be worthwhile. Clinical profiling could be improved in future studies by including additional dedicated assessments, such as a neuropsychological battery, the Unified Parkinson’s Disease Rating Scale (UPDRS), modified PSPRS and specific cerebellar scales. Whilst it remains to be proven, molecular and microstructural imaging methods may add to our recognition of cerebellar involvement before the emergence of significant atrophy.

Future dedicated studies of the cerebellum using these methods, would potentially improve detection of significant clinico-radiologic correlations. Lastly, longitudinal studies are needed to better understand the evolution of cerebellar changes in PSP. Improving our understanding of the cerebellum in PSP, potentially contributes to our interpretation and application of *in vivo* imaging in clinical practice. Furthermore, in time, we may find that cerebellar changes predominate in certain variant presentations, though this requires further study.

There are several strengths and limitations of our study. First, PSP is a rare condition, so our sample size of thirty-seven is larger than in many prior studies of this disease. Additionally, we have used optimised automated MRI pipelines to objectively assess subtentorial brain regions, which enables accurate individualised estimation of regional volumes. We chose to limit the number of ROIs included in this study in order to limit the extent of multiple comparison corrections.

However, this also limited the anatomical specificity of our inferences, including examination of smaller, lobule-level ROIs and investigations of asymmetric changes, which could shed more light on disease morphology. An exploratory analysis could be considered in this regard; however, the interpretation would be limited due to the modest sample size.

International, multisite studies that aggregate MRI data from a large number of subjects, which have been successful in this regard in other rare neurodegenerative diseases^39^, may provide one way forward.

Lastly, heterogeneity within our cohort is an important limitation. While most PSP patients have probable PSP-RS, we included individuals with several additional variants in the analysis. These are known to present differently, with varying rates and distributions of tau spread, despite core pathology in the basal ganglia and midbrain^1,4^. Statistical comparisons between variants could not be made to further elucidate these differences due to the small number, but box plots with variants are shown in **Supplementary Figure 1**, and we have largely replicated the results from the main analysis when restricting the analysis only to our predominate group of patients with PSP-RS **(Supplementary Table 1)**.

## Conclusion

Alongside midbrain atrophy, this study demonstrates volume loss in additional brainstem and cerebellar regions in PSP. Most importantly, these comprise several cerebellar regions, including the flocculonodular lobe and cerebellar white matter, highlighting that the cerebellum is not entirely spared in this disease. Moreover, important clinico-radiologic correlations were detected which suggest a crucial role of the cerebellum and cerebellar connections in non-motor and motor PSP symptoms. Most notably, cognitive dysfunction in PSP was shown to correlate with the superior and inferior posterior lobe volume, and PSP disease severity with superior cerebellar peduncle volume. These findings, alongside consistent trends indicating the potential for more widespread involvement of cerebellar cortical regions, motivates the need for further research to clarify the contribution of the cerebellum to interindividual symptom variability, and the need to incorporate the cerebellum into PSP disease models.

## Acknowledgements

Author roles:

(1) Research Project: A. Design, B. Organization, C. Execution; (2) Analysis: A. Design, B. Execution, C. Review and Critique; (3) Manuscript Preparation: A. Writing of the First Draft, B. Review and Critique.

CS: 1A, 1B, 1C, 2A, 2B, 2C, 3A, 3B

TS: 1A, 1B, 1C, 2B

CM: 2A, 3B

JJL: 1C, 3B

KB: 1A, 3B

TOB: 1A, 3B

ML: 1B, 2B

LV: 1A, 2A, 2C, 3B

IHH: 1A, 2A, 2C, 3B

## Financial disclosures

CS, TS, CM receive an Australian Government Research Training Program Stipend. TOB’s institution has received research funding and consultancy fees from UCB, Eisai, LivaNova, Supernus, Praxis Pharmaceuticals, ES Therapeutics and Biogen, outside the submitted work. LV reports funding from NHMRC Medical Research Future Fund (MRF1170276, MRF1200254, MRF2023250), Multiple Sclerosis Research Association, and Department of Science Industry and Resources for unrelated projects. IHH receives grants unrelated to this work from the Australian National Health and Medical Research Council and the Friedreich Ataxia Research Alliance, and is a consultant for Solid Biosciences Inc.

## Supplementary Material

**Supplementary Figure 1.**
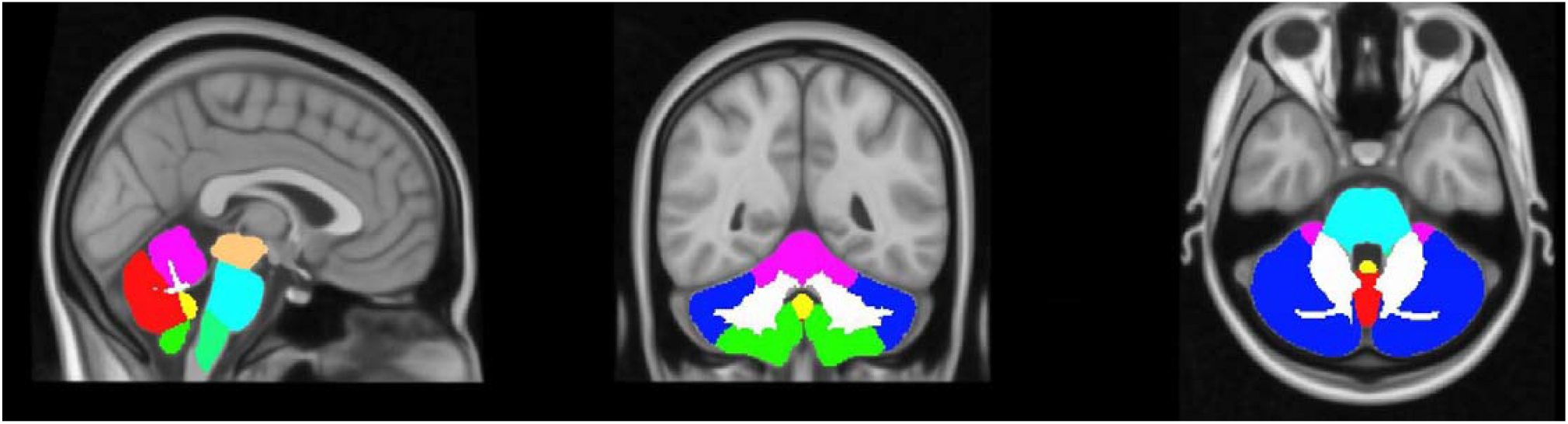
Brainstem and cerebellar regions of interest. Brainstem and cerebellar regions of interest in colour segments: Midbrain (orange), pons (light blue), medulla (green), anterior lobe (purple), superior posterior lobe (dark blue), vermis (red) flocculonodular lobe (yellow) and inferior posterior lobe (green), corpus medullare (white). *LEFT = sagittal view in midline CENTRE = coronal view showing cerebellar segments RIGHT = axial view at the pons level*.

**Supplementary table 1.**
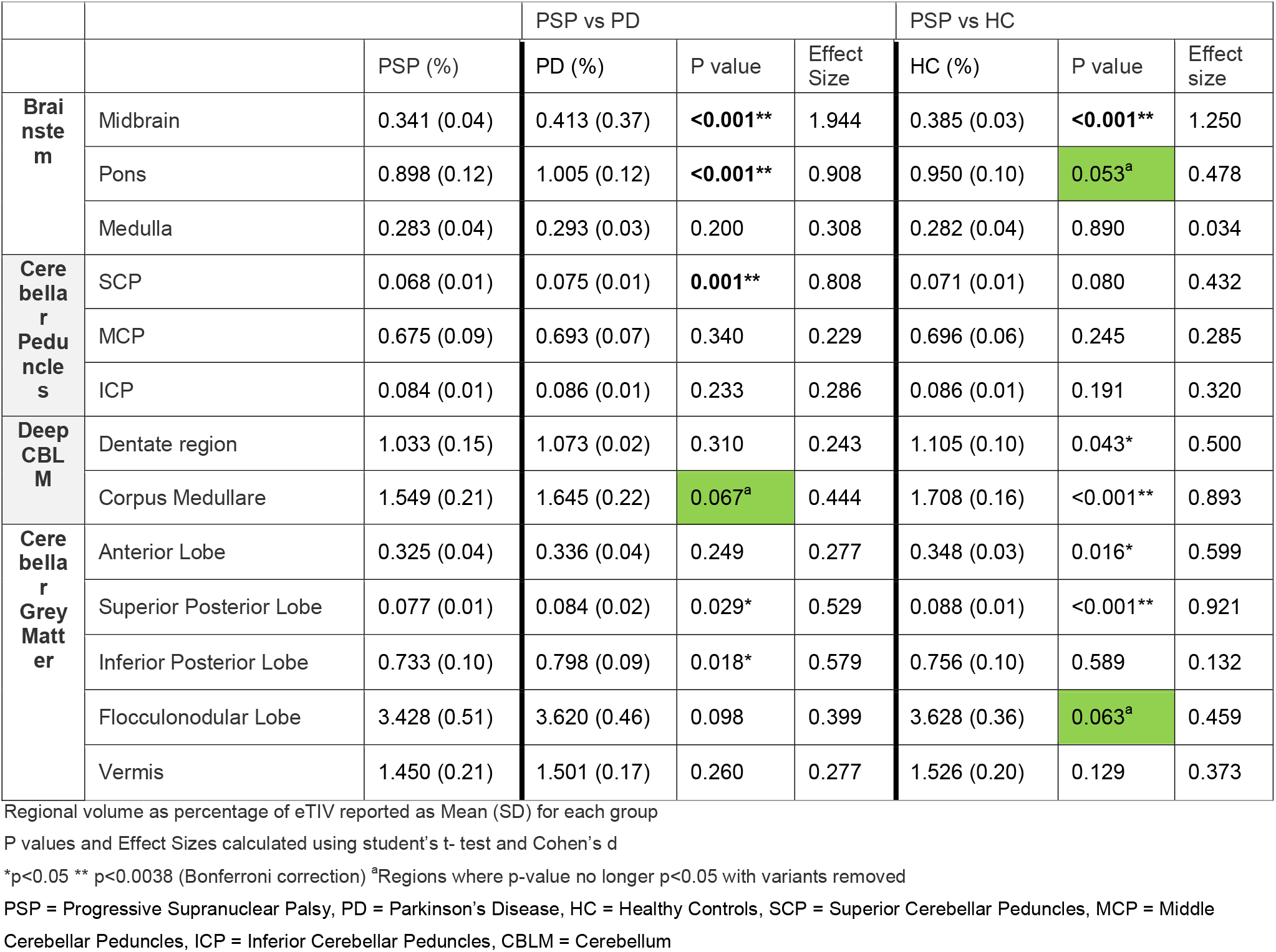
Comparisons of regional volumetric findings for PSP-RS only

## Notes

**Conflict of interest** statement The authors declare that there are no competing financial or personal interests that could have influenced this work.

### Competing Interest Statement

The authors have declared no competing interest.

### Funding Statement

This study was funded by a Medical Research Future Fund grant from the Australian Government to TOB and LV (MRF1200254).

### Author Declarations

Ethics approval was obtained from the Human Research Ethics Committee at the Alfred Hospital (Monash-Alfred atypical parkinsonian database project number 157/19; SEL003 project number 594/20; ACTRN12600001254987). Written informed consent was obtained from all participants recruited.

## References

1. Kovacs GG, Lukic MJ, Irwin DJ, et al. Distribution patterns of tau pathology in progressive supranuclear palsy. Acta Neuropathol (Berl). 2020;140(2):99–119. doi:10.1007/s00401-020-02158-2

2. Höglinger GU, Respondek G, Stamelou M, et al. Clinical diagnosis of progressive supranuclear palsy: The movement disorder society criteria: MDS Clinical Diagnostic Criteria for PSP. Mov Disord. 2017;32(6):853–864. doi:10.1002/mds.26987

3. Whitwell JL, Höglinger GU, Antonini A, et al. Radiological biomarkers for diagnosis in PSP: Where are we and where do we need to be?: Neuroimaging Biomarkers for Diagnosis in PSP. Mov Disord. 2017;32(7):955–971. doi:10.1002/mds.27038

4. Williams DR, Holton JL, Strand C, et al. Pathological tau burden and distribution distinguishes progressive supranuclear palsy-parkinsonism from Richardson’s syndrome. Brain. 2007;130(6):1566–1576. doi:10.1093/brain/awm104

5. Chatterjee K, Paul S, Banerjee R, et al. Characterizing gait and exploring neuro-morphometry in patients with PSP-Richardson’s syndrome and vascular parkinsonism. Park Relat Disord. 2023;113:105483. doi:10.1016/j.parkreldis.2023.105483

6. Tse NY, Chen Y, Irish M, et al. Cerebellar contributions to cognition in corticobasal syndrome and progressive supranuclear palsy. Brain Commun. 2020;2(2):fcaa194. doi:10.1093/braincomms/fcaa194

7. Anagnostou E., Karavasilis E., Potiri I., et al. A cortical substrate for square-wave jerks in progressive supranuclear palsy. J Clin Neurol Korea. 2020;16(1):37–45. doi:10.3988/jcn.2020.16.1.37

8. Seki M., Seppi K., Mueller C., et al. Diagnostic potential of dentatorubrothalamic tract analysis in progressive supranuclear palsy. Parkinsonism Relat Disord. 2018;49:81–87. doi:10.1016/j.parkreldis.2018.02.004

9. Paviour D., Price S.L., Jahanshahi M., Lees A.J., Fox N.C. Regional brain volumes distinguish PSP, MSA-P, and PD: MRI-based clinico-radiological correlations. Mov Disord. 2006;21(7):989–996. doi:10.1002/mds.20877

10. Giordano A., Tessitore A., Corbo D., et al. Clinical and cognitive correlations of regional gray matter atrophy in progressive supranuclear palsy. Parkinsonism Relat Disord. 2013;19(6):590–594. doi:10.1016/j.parkreldis.2013.02.005

11. Agosta F, Kostic VS, Galantucci S, et al. The in vivo distribution of brain tissue loss in Richardson’s syndrome and PSP-parkinsonism: a VBM-DARTEL study. Eur J Neurosci. 2010;32(4):640–647. doi:10.1111/j.1460-9568.2010.07304.x

12. Pan P., Liu Y., Zhang Y., Zhao H., Ye X., Xu Y. Brain gray matter abnormalities in progressive supranuclear palsy revisited. Oncotarget. 2017;8(46):80941–80955. doi:10.18632/oncotarget.20895

13. Gellersen HM, Guo CC, O’Callaghan C, Tan RH, Sami S, Hornberger M. Cerebellar atrophy in neurodegeneration—a meta-analysis. J Neurol Neurosurg Psychiatry. 2017;88(9):780–788. doi:10.1136/jnnp-2017-315607

14. Yu F., Barron D.S., Tantiwongkosi B., Fox P. Patterns of gray matter atrophy in atypical parkinsonism syndromes: A VBM meta-analysis. Brain Behav. 2015;5(6):1–10. doi:10.1002/brb3.329

15. Shao N., Yang J., Li J., Shang H.-F. Voxelwise meta-analysis of gray matter anomalies in progressive supranuclear palsy and Parkinson’s disease using anatomic likelihood estimation. Front Hum Neurosci. 2014;8(1 FEB):63. doi:10.3389/fnhum.2014.00063

16. Lewis MM, Galley S, Johnson S, Stevenson J, Huang X, McKeown MJ. The Role of the Cerebellum in the Pathophysiology of Parkinson’s Disease. Can J Neurol Sci J Can Sci Neurol. 2013;40(3):299–306. doi:10.1017/S0317167100014232

17. McDonald CR, Hagler DJJ, Ahmadi ME, et al. Subcortical and cerebellar atrophy in mesial temporal lobe epilepsy revealed by automatic segmentation. Epilepsy Res. 2008;79(2-3):130–138. doi:10.1016/j.eplepsyres.2008.01.006

18. Möller C, Vrenken H, Jiskoot L, et al. Different patterns of gray matter atrophy in early- and late-onset Alzheimer’s disease. Neurobiol Aging. 2013;34(8):2014–2022. doi:10.1016/j.neurobiolaging.2013.02.013

19. Siejka TP, Bertram KL, Tang HM, et al. Monash-Alfred protocol for assessment of atypical parkinsonian syndromes (MAP-APS). BMJ Neurol Open. 2024;6(1):e000553. doi:10.1136/bmjno-2023-000553

20. Faber J, Kügler D, Bahrami E, et al. CerebNet: A fast and reliable deep-learning pipeline for detailed cerebellum sub-segmentation. NeuroImage. 2022;264:119703. doi:10.1016/j.neuroimage.2022.119703

21. Oishi K, Faria A, Jiang H, et al. Atlas-based whole brain white matter analysis using large deformation diffeomorphic metric mapping: application to normal elderly and Alzheimer’s disease participants. NeuroImage. 2009;46(2):486–499. doi:10.1016/j.neuroimage.2009.01.002

22. Diedrichsen J, Maderwald S, Küper M, et al. Imaging the deep cerebellar nuclei: a probabilistic atlas and normalization procedure. NeuroImage. 2011;54(3):1786–1794. doi:10.1016/j.neuroimage.2010.10.035

23. Cosottini M, Ceravolo R, Faggioni L, et al. Assessment of midbrain atrophy in patients with progressive supranuclear palsy with routine magnetic resonance imaging. Acta Neurol Scand. 2007;116(1):37–42. doi:10.1111/j.1600-0404.2006.00767.x

24. Whitwell J.L., Tosakulwong N., Botha H., et al. Brain volume and flortaucipir analysis of progressive supranuclear palsy clinical variants. NeuroImage Clin. 2020;25:102152. doi:10.1016/j.nicl.2019.102152

25. Sjostrom H, Granberg T, Hashim F, Westman E, Svenningsson P. Automated brainstem volumetry can aid in the diagnostics of parkinsonian disorders. Parkinsonism Relat Disord. 2020;79(9513583):18–25. doi:10.1016/j.parkreldis.2020.08.004

26. Mueller K., Jech R., Bonnet C., et al. Disease-specific regions outperform whole-brain approaches in identifying progressive supranuclear palsy: A multicentric MRI study. Front Neurosci. 2017;11(MAR):100. doi:10.3389/fnins.2017.00100

27. Scherfler C., Gobel G., Muller C., et al. Diagnostic potential of automated subcortical volume segmentation in atypical parkinsonism. Neurology. 2016;86(13):1242–1249. doi:10.1212/WNL.0000000000002518

28. Albrecht F, Bisenius S, Neumann J, Whitwell J, Schroeter ML. Atrophy in midbrain & cerebral/cerebellar pedunculi is characteristic for progressive supranuclear palsy - A double-validation whole-brain meta-analysis. NeuroImage Clin. 2019;22:101722. doi:10.1016/j.nicl.2019.101722

29. Matsuoka K., Takado Y., Tagai K., et al. Two pathways differentially linking tau depositions, oxidative stress, and neuronal loss to apathetic phenotypes in progressive supranuclear palsy. J Neurol Sci. 2023;444:120514. doi:10.1016/j.jns.2022.120514

30. Woo K.A., Shin J.Y., Kim H., Ahn J., Jeon B., Lee J.-Y. Peripapillary retinal nerve fiber layer thinning in patients with progressive supranuclear palsy. J Neurol. 2022;269(6):3216–3225. doi:10.1007/s00415-021-10936-5

31. Potrusil T., Krismer F., Beliveau V., et al. Diagnostic potential of automated tractography in progressive supranuclear palsy variants. Parkinsonism Relat Disord. 2020;72:65–71. doi:10.1016/j.parkreldis.2020.02.007

32. Sintini I., Schwarz C.G., Senjem M.L., et al. Multimodal neuroimaging relationships in progressive supranuclear palsy. Parkinsonism Relat Disord. 2019;66:56–61. doi:10.1016/j.parkreldis.2019.07.001

33. Dutt S., Binney R.J., Heuer H.W., et al. Progression of brain atrophy in PSP and CBS over 6 months and 1 year. Neurology. 2016;87(19):2016–2025. doi:10.1212/WNL.0000000000003305

34. Focke N.K., Helms G., Scheewe S., et al. Individual voxel-based subtype prediction can differentiate progressive supranuclear palsy from idiopathic Parkinson syndrome and healthy controls. Hum Brain Mapp. 2011;32(11):1905–1915. doi:10.1002/hbm.21161

35. Abos A., Segura B., Baggio H.C., et al. Disrupted structural connectivity of fronto-deep gray matter pathways in progressive supranuclear palsy. NeuroImage Clin. 2019;23:101899. doi:10.1016/j.nicl.2019.101899

36. Schonhaut D.R., McMillan C.T., Spina S., et al. 18F-flortaucipir tau positron emission tomography distinguishes established progressive supranuclear palsy from controls and Parkinson disease: A multicenter study. Ann Neurol. 2017;82(4):622–634. doi:10.1002/ana.25060

37. Surova Y., Nilsson M., Latt J., et al. Disease-specific structural changes in thalamus and dentatorubrothalamic tract in progressive supranuclear palsy. Neuroradiology. 2015;57(11):1079–1091. doi:10.1007/s00234-015-1563-z

38. Stoodley CJ, Schmahmann JD. Functional topography of the human cerebellum. Handb Clin Neurol. 2018;154:59–70. doi:10.1016/B978-0-444-63956-1.00004-7

39. Harding IH, Chopra S, Arrigoni F, et al. Brain Structure and Degeneration Staging in Friedreich Ataxia: Magnetic Resonance Imaging Volumetrics from the ENIGMA-Ataxia Working Group. Ann Neurol. 2021;90(4):570–583. doi:10.1002/ana.26200

