## Supplementary Materials for "Brainstem and cerebellar volume loss and the associated clinical features in Progressive Supranuclear Palsy"

**Supplementary Material**

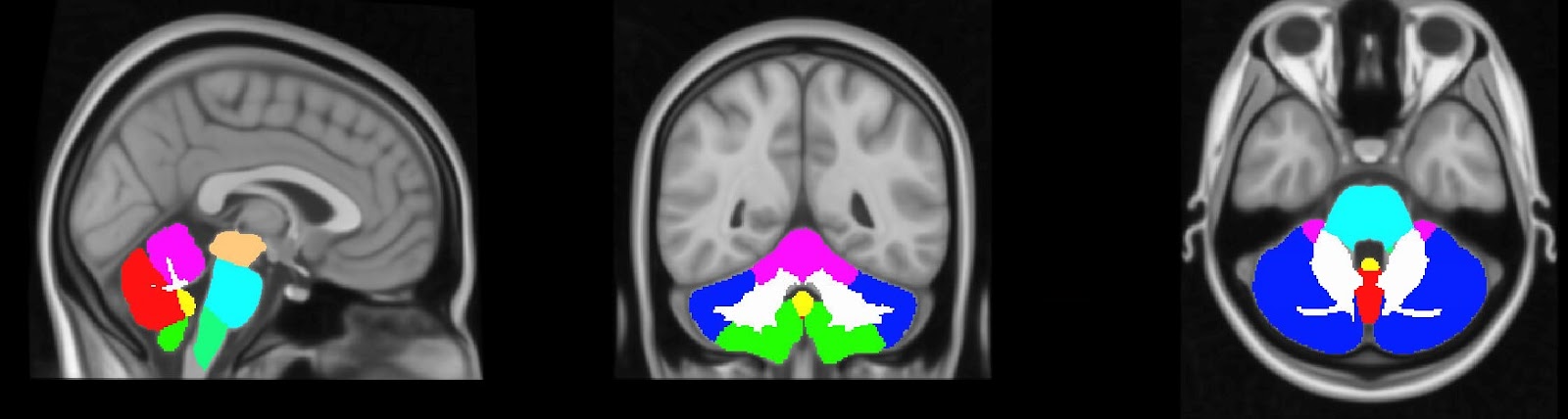

**Supplementary Figure 1. Brainstem and cerebellar regions of interest**

Brainstem and cerebellar regions of interest in colour segments: Midbrain (orange), pons (light blue), medulla (green), anterior lobe (purple), superior posterior lobe (dark blue), vermis (red) flocculonodular lobe (yellow) and inferior posterior lobe (green), corpus medullare (white). *LEFT = sagittal view in midline CENTRE = coronal view showing cerebellar segments RIGHT = axial view at the pons level.*

**Supplementary table 1. Comparisons of regional volumetric findings for PSP-RS only**

|  |  | | PSP vs PD | | | PSP vs HC | | |
| --- | --- | --- | --- | --- | --- | --- | --- | --- |
|  |  | PSP (%) | PD (%) | P value | Effect Size | HC (%) | P value | Effect size |
| **Brainstem** | Midbrain | 0.341 (0.04) | 0.413 (0.37) | **<0.001**** | 1.944 | 0.385 (0.03) | **<0.001**** | 1.250 |
|  | Pons | 0.898 (0.12) | 1.005 (0.12) | **<0.001**** | 0.908 | 0.950 (0.10) | 0.053^a^ | 0.478 |
|  | Medulla | 0.283 (0.04) | 0.293 (0.03) | 0.200 | 0.308 | 0.282 (0.04) | 0.890 | 0.034 |
| **Cerebellar Peduncles** | SCP | 0.068 (0.01) | 0.075 (0.01) | **0.001**** | 0.808 | 0.071 (0.01) | 0.080 | 0.432 |
|  | MCP | 0.675 (0.09) | 0.693 (0.07) | 0.340 | 0.229 | 0.696 (0.06) | 0.245 | 0.285 |
|  | ICP | 0.084 (0.01) | 0.086 (0.01) | 0.233 | 0.286 | 0.086 (0.01) | 0.191 | 0.320 |
| **Deep CBLM** | Dentate region | 1.033 (0.15) | 1.073 (0.02) | 0.310 | 0.243 | 1.105 (0.10) | 0.043* | 0.500 |
|  | Corpus Medullare | 1.549 (0.21) | 1.645 (0.22) | 0.067^a^ | 0.444 | 1.708 (0.16) | <0.001** | 0.893 |
| **Cerebellar Grey Matter** | Anterior Lobe | 0.325 (0.04) | 0.336 (0.04) | 0.249 | 0.277 | 0.348 (0.03) | 0.016* | 0.599 |
|  | Superior Posterior Lobe | 0.077 (0.01) | 0.084 (0.02) | 0.029* | 0.529 | 0.088 (0.01) | <0.001** | 0.921 |
|  | Inferior Posterior Lobe | 0.733 (0.10) | 0.798 (0.09) | 0.018* | 0.579 | 0.756 (0.10) | 0.589 | 0.132 |
|  | Flocculonodular Lobe | 3.428 (0.51) | 3.620 (0.46) | 0.098 | 0.399 | 3.628 (0.36) | 0.063^a^ | 0.459 |
|  | Vermis | 1.450 (0.21) | 1.501 (0.17) | 0.260 | 0.277 | 1.526 (0.20) | 0.129 | 0.373 |

Regional volume as percentage of eTIV reported as Mean (SD) for each group

P values and Effect Sizes calculated using student’s t- test and Cohen’s d

*p<0.05 ** p<0.0038 (Bonferroni correction) ^a^Regions where p-value no longer p<0.05 with variants removed

PSP = Progressive Supranuclear Palsy, PD = Parkinson’s Disease, HC = Healthy Controls, SCP = Superior Cerebellar Peduncles, MCP = Middle Cerebellar Peduncles, ICP = Inferior Cerebellar Peduncles, CBLM = Cerebellum
